# Quantitative Magnetic Resonance Cerebral Spinal Fluid Flow Properties and Executive Function Cognitive Outcomes in Congenital Heart Disease

**DOI:** 10.1101/2024.04.19.24306104

**Authors:** Vincent Kyu Lee, William T. Reynolds, Julia Wallace, Nancy Beluk, Daryaneh Badaly, Cecilia W Lo, Rafael Ceschin, Ashok Panigrahy

## Abstract

Cerebrospinal fluid (CSF) circulation has recently been shown to be important in nutrient distribution, waste removal, and neurogenesis. Increased CSF volumes are frequently observed in congenital heart disease (CHD) and are associated with neurodevelopmental deficits. This suggests prolonged perturbation to the CSF system and possible interference to its homeostatic function, which may contribute to the neurodevelopmental deficits in CHD. CSF flow has yet to be studied in CHD patients, but the pulsatile flow of CSF throughout the brain is driven mainly by cardiopulmonary circulation. Given the underlying heart defects in CHD, the cardiopulmonary circulatory mechanisms in CHD might be impaired with resultant perturbation on the CSF circulation. In this study, we determine whether CSF flow, using MRI measurements of static and dynamic pulsatile flow, is abnormal in youths with CHD compared to healthy controls in relation to executive cognitive function. CSF flow measurements were obtained on a total of 58 child and young adult participants (CHD=20, healthy controls = 38). The CSF flow was measured across the lumen of the Aqueduct of Sylvius using cardiac-gated phase-contrast MRI at 3.0T. Static pulsatility was characterized as anterograde and retrograde peak velocities, mean velocity, velocity variance measurements, and dynamic pulsatility calculated as each participant’s CSF flow deviation from the study cohort’s consensus flow measured with root mean squared deviation (RMSD) were obtained. The participants had neurocognitive assessments for executive function with focus on inhibition, cognitive flexibility, and working memory domains. The CHD group demonstrated greater dynamic pulsatility (higher overall flow RMSD over the entire CSF flow cycle) compared to controls (p=0.0353), with no difference detected in static pulsatility measures. However, lower static CSF flow pulsatility (anterograde peak velocity: p=0.0323) and lower dynamic CSF flow pulsatility (RMSD: p=0.0181) predicted poor inhibitory executive function outcome. Taken together, while the whole CHD group exhibited higher dynamic CSF flow pulsatility compared to controls, the subset of CHD subjects with relatively reduced static and dynamic CSF flow pulsatility had the worst executive functioning, specifically the inhibition domain. These findings suggest that altered CSF flow pulsatility may be central to not only brain compensatory mechanisms but can also drive cognitive impairment in CHD. Further studies are needed to investigate possible mechanistic etiologies of aberrant CSF pulsatility (i.e. primary cardiac hemodynamic disturbances, intrinsic brain vascular stiffness, altered visco-elastic properties of tissue, or glial-lymphatic disturbances), which can result in acquired small vessel brain injury (including microbleeds and white matter hyperintensities).

## Introduction

Abnormal cerebrospinal fluid (CSF) findings, especially increased CSF volume, are frequently observed in studies of brain development in the context of congenital heart disease (CHD).[1–5] Extra-axial CSF have been observed in complex CHD neonates and even in third trimester fetuses.[6–9] Increased CSF volume has been shown to be correlated with brain dysmaturation (i.e., dysplastic cortical and subcortical structures, decreased cortical and subcortical volumes, and decreased cortical thickness), as well as poor neurodevelopmental outcomes in preoperative neonates[10] and young children with CHD.[2,3,11]

It is currently unknown if there are CSF flow disturbances in CHD, extending the spectrum of the type of CSF phenotypic abnormalities in CHD patients. The CSF circulation in conjunction with the glymphatic system is shown to play an important role in nutrient distribution, waste removal, and neurogenesis.[12,13] Prolonged impairment of this important homeostatic function may lead to neuroinflammation and neurodevelopmental deficits.[14] In vivo CSF flow studies in humans have just begun recently with the advent of phase contrast magnetic resonance imaging (MRI). While much remains to be characterized, the flow of CSF through the ventricles, cisterns, and perivascular spaces of the brain are pulsatile, driven mainly by cardiopulmonary circulation[15] with arterial pulsations as one of the primary drivers.[16] This mechanism is likely impaired in CHD, and there may be considerably increased demands on the CSF circulation to clear toxic and inflammatory molecules, especially for heart defects that result in hypoxia and cyanosis. Therefore, understanding the presence of differences in CSF flow in CHD, beyond differences in CSF volume, is critical.

Given the etiology of heart defects and subsequent repairs, this circulatory mechanism in CHD might be impaired with resultant perturbation on the CSF circulation, especially in complex heart defects such as hypoplastic left heart syndrome (HLHS) that result in hypoxia and cyanosis. As a consequence, there may be considerably increased demands on the CSF circulation to clear toxic and inflammatory molecules, especially for heart defects that result in hypoxia and cyanosis. While not in CHD patients, CSF abnormalities have been linked to autism spectrum disorder in early childhood MRI studies of pediatric ASD[17]. Abnormal CSF flow has also been shown, albeit in elderly patients, to be associated with cognitive decline[18]. These studies collectively suggest that further delineation of CSF properties may provide mechanistic insight in relation to understanding adverse neurodevelopmental outcomes in children with CHD.

Here, based on prior flow studies [4], we utilized MRI phase contrast imaging (PCMRI) at the level of the aqueduct of Sylvius in the midbrain to study CSF flow properties in youths with CHD for the first time compared to healthy controls. We also assessed whether these CSF flow properties predicted poor neurocognitive function (focusing on executive function) in CHD. Given the frequent occurrence of increased CSF volume in CHD, we also correlated CSF volume (from different anatomic portions of the ventricular system) with these CSF flow properties. While the primary focus of our analysis was based on traditional static CSF flow pulsatility metrics we secondarily developed an innovative quantitative method to capture the dynamic variation of CSF flow pulsatility over the entire cardiac cycle.

## Material and Methods

### Participants

Participants were recruited from a single center, using print and digital advertisements, an online registry of healthy volunteers, and referrals within targeted cardiology outpatient clinics. Study exclusion criteria included comorbid genetic disorders, contraindications for MRI (e.g., a pacemaker), and non-English speakers. For healthy controls, study exclusion criteria also included preterm birth and neurological abnormalities (e.g., brain malformations, strokes, hydrocephalus). We initially screened 143 patients with CHD and 98 healthy controls. A total of 69 CHD and 92 healthy controls underwent brain MR scanning. The MRI CSF phase contrast flow sequences were added to the end of the imaging protocol for a subset of patients nested within this larger cohort including 26 CHD patients and 49 healthy controls. After removing cases with un-analyzable imaging due to motion artifacts or technical factors, the final sample of patients included the following: CHD patients (n=20) and age-matched controls (n=38). These patients were prospectively recruited at our institution with Institutional Review Board (IRB) approval and oversight (for reference: University of Pittsburgh Institutional Review Board STUDY20060128: Multimodal Connectome Study approval 23 July 2020 and STUDY1904003 Ciliary Dysfunction, Brain Dysplasia, and Neurodevelopmental Outcome in Congenital approval 6 February 2023). The project was completed in accordance with the ethical principles of the Helsinki Declaration. We have previously published portions of this prospectively recruited cohort.[19–23] Patients with CHD included a heterogenous mix of cardiac lesions, including hypoplastic left heart syndrome (HLHS), aortic arch abnormalities, d-transposition of the great aorta (d-TGA), and other malformations requiring surgical correction in the first year of life. Clinical and surgical history from birth, medical risk factors, and additional social determinants of health (SDOH) were collected from the medical record.

### Imaging

Participants were scanned with an Institutional Review Board approved MRI protocol on the same Siemens 3T Skyra magnet (Siemens Healthcare, Erlangen, Germany) using a 32-channel head coil. The CSF flow portion of the exam was acquired using phase contrast gradient echo imaging sequence with cardiac-gated trigger. This through plane flow measurement was acquired with a transversal slice placed perpendicularly across the level of the Aqueduct of Silvius (Figure 1A) with the following parameters: velocity encoding = ±12 cm/s; TR/TE = 9.66/30.40 ms; flip angle = 15°; matrix = 256 x 256; in plane resolution of 0.6mm x 0.6mm; slice thickness = 5mm, averaged over 150 repeated measures. The participants also received a volumetric T1 scan acquired in sagittal orientation with MPRAGE sequence using the following parameters: TE/TR=3.2/2400 ms; matrix=256×256; resolution 1.0×1.0×1.0 mm^3^.

**Figure 1.**
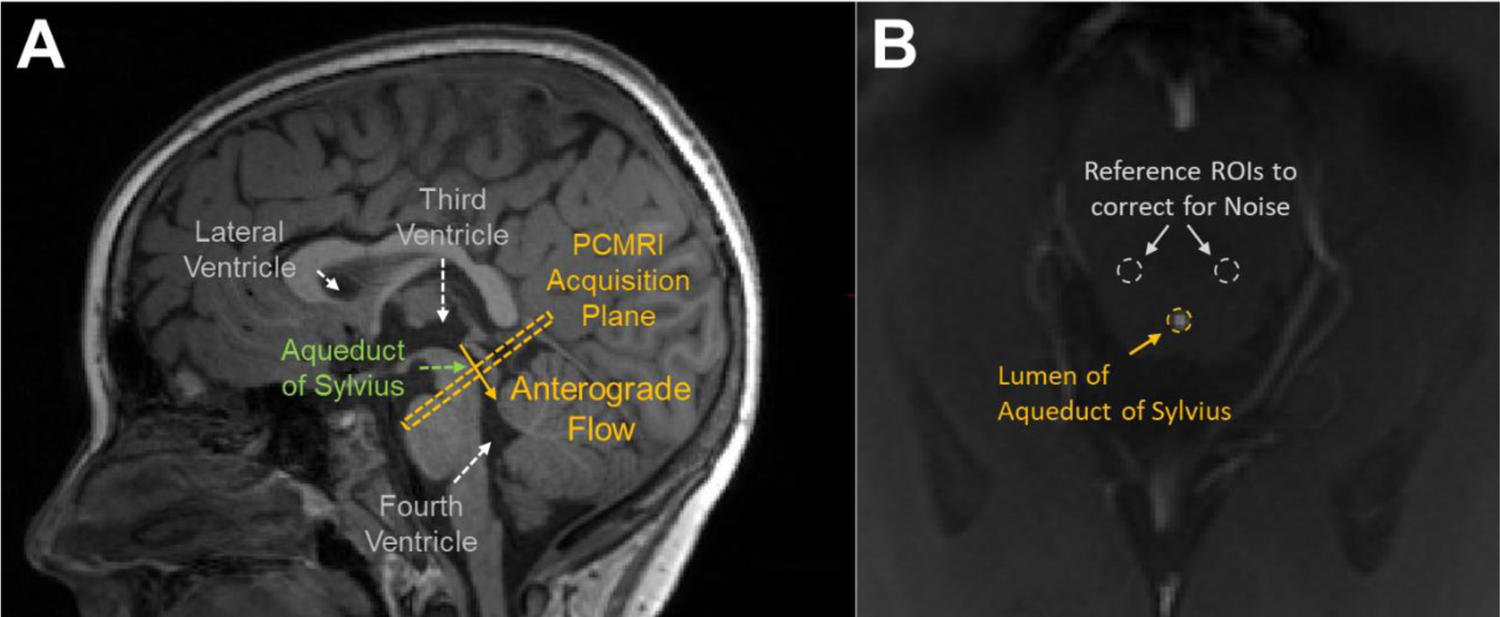
(A) The CSF flow was measured across the lumen of the Aqueduct of Sylvius using a transverse slice of 5mm thickness (yellow dashed rectangle). The physiologically forward or down-stream flow direction of CSF was set as the anterograde flow direction (yellow solid arrow). (B) The in-plane cross-sectional image of the brain at the level of the Aqueduct, showing the ROIs used to measure the flow velocities in the lumen of the aqueduct (yellow dotted circle) and the reference measurements in surrounding brain parenchyma to correct for noise (white dotted circles).

### CSF Flow Measurement

PCMRI acquisition is cardiac-gated and 20 images at regularly spaced time intervals are sampled over the cardiac cycle. The flow dynamic and metrics of the aqueduct for each participant are calculated from the magnitude and phase image acquisition using a processing pipeline script programmed in MATLAB. For each set of CSF images, a region of interest (ROI) encompassing the lumen of the aqueduct was used. To correct for noise, two similarly sized ROIs placed on either side of the aqueduct in the surrounding brain parenchyma was used as reference to measure noise (Figure 1B).

For consistent and complete coverage of the lumen, the mask fit was assessed at all PCMRI slices, with adjustments made to ROI as necessary. The forward direction of CSF flow in this study was oriented following the physiological flow of CSF, which is from the third ventricle to the fourth ventricle. Thus, the anterograde direction is set as the forward or down stream flow of CSF through the Aqueduct of Sylvius from third ventricle to fourth ventricle (yellow arrow Figure 1A). Conversely, the retrograde direction is set as flow of CSF from fourth to third ventricle.

Figure 2 illustrates the CSF flow characteristics and features measured from each set of PCMRI data, and they are as follows. CSF flow velocity encompass the rates of the fluid flowing through the lumen and are measured in cm/sec. Anterograde peak velocity (APVel) represents the greatest magnitude of velocity measured in the anterograde direction defined as flowing from third to fourth ventricle. Retrograde peak velocity (RPVel) represents the greatest magnitude of velocity measured in the retrograde direction defined as flowing from the fourth to the third ventricle. Mean Velocity (MVel) is calculated from mean of all velocity measurements over the 20 sample points, and the square of the standard deviation of these velocities is the Velocity Variance (VVel).

**Figure 2.**
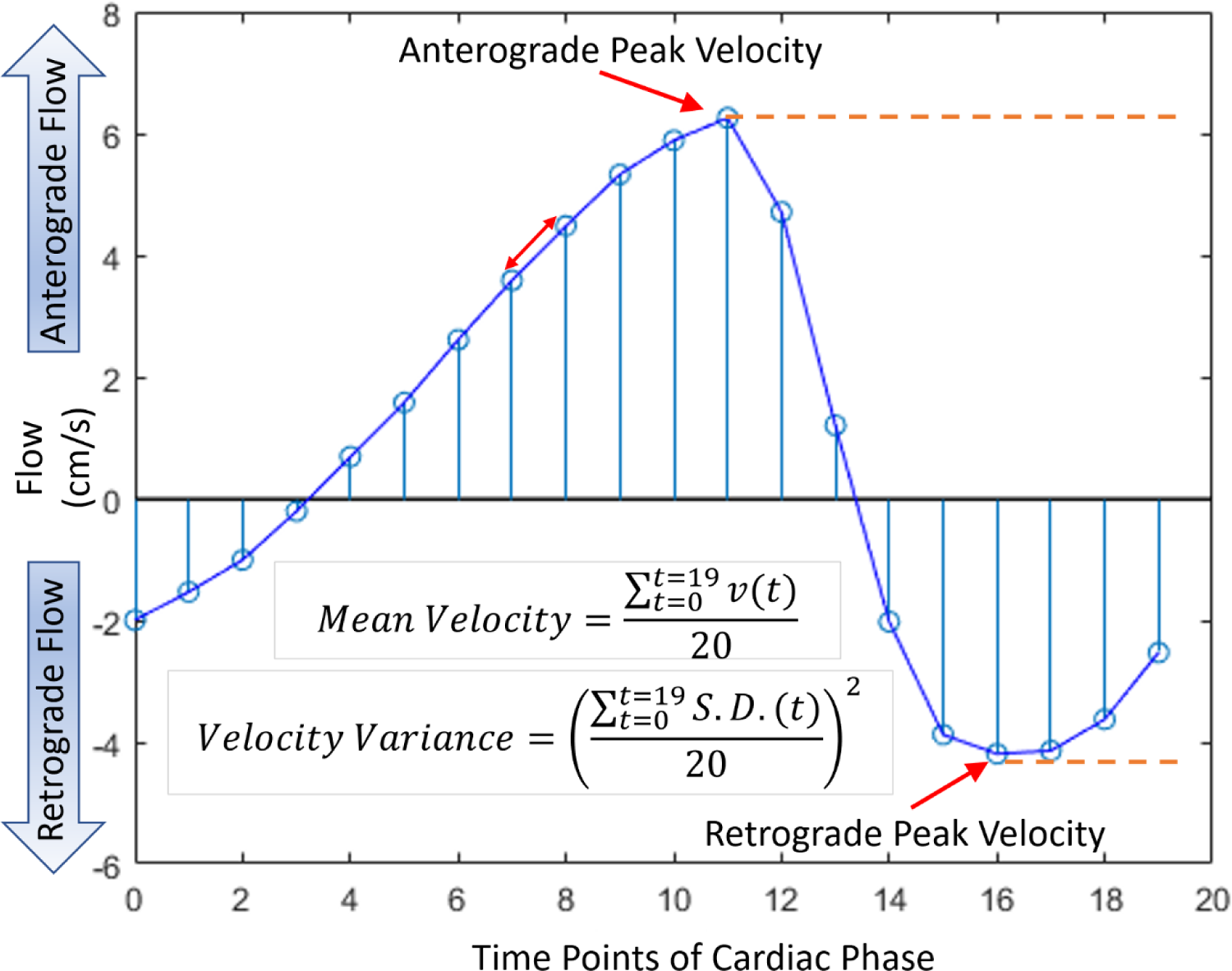
Feature extraction diagram for standard and traditional flow measurements: anterograde peal velocity (APVel) and retrograde peak velocity (RPVel), mean velocity over the entire period (MVel). The velocity variance (VVel) is the squared standard deviations of the flow at each time point.

### CSF flow variance over the entire pulsatile CSF flow cycle

CSF flow can be characterized as the various features described in the previous section. However, these features, by their nature, represent isolated or lone aspects of CSF flow. No inferential connections can be made between these features analytically. No inferential connections can be made about the CSF flow over its pulsatile cycles just using these features.

However, CSF flow is a continuous phenomenon that is likely related to cardiac pulsations and intertwined with the cardiac cycle. The standard CSF flow measurements of velocities and volumes do not encapsulate the multidimensional nature of the CSF flow in its entirety. When peak velocities or net flow volumes are compared between individuals, we are only comparing lone aspects of the CSF flow, and not seeing the overall flow similarity or differences between these participants.

To characterize the CSF flow variance as a pulsatile phenomenon over the entire cardiac cycle, measurements over the entirety of the PCMRI series were evaluated as illustrated in Figure 3. The PCMRI data of the entire cohort was used to fit a sinusoidal function representing the consensus CSF flow (Figure 3A). This fitted curve represents the group average or norm. Consequently, the deviation and difference of each participant’s CSF flow characteristic over the entirety of the cardiac cycle can be determined from this group norm fitted curve by quantifying the variance of each person’s flow curve using root mean squared deviation (RMSD) calculations (Figure 3B). This essentially measures the degree of similarity or difference of a person’s CSF flow to the group consensus average as a RMSD value with higher values indicating greater difference.

**Figure 3.**
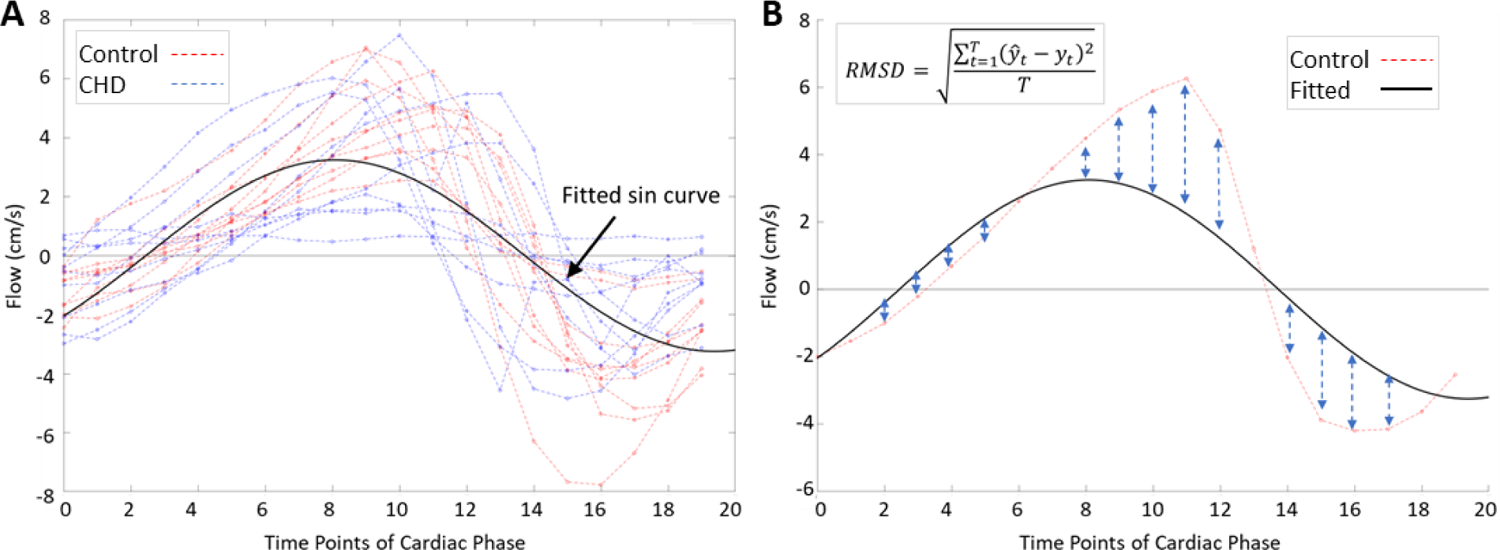
(A) Evaluation of the CSF Flow in its entirety over the cardiac cycle. Using the PCMRI data of the entire cohort to fit a sin curve, and (B) quantify the difference of each individual’s PCMRI data from the cohort’s fitted sin curve.

### CSF Volume Segmentation

CSF volume measurements were generated from the T1 images. For the intra-ventricular CSF measurements, standard FreeSurfer software volume parcellations was used[24,25]. Based on the parcellation, the ventricular compartment was divided as follows: left and right lateral ventricles, left and right inferior lateral ventricles, third ventricle, and fourth ventricle. The whole brain CSF was determined using FSL’s FAST segmentation[26] method, and the extra axial CSF volume was calculated from the difference between this total whole brain CSF volume and intraventricular volumes.

### Cognitive Assessment

The study participants received a battery of cognitive tests, and ratings of their cognitive functioning were collected. Within the limited scope of this study, we focused on tests od executive function, particularly cognitive flexibility, inhibitory control, and working memory (i.e., core executive functions based on the model of Diamond, 2013).[27] Participants completed measures from the National Institute of Health Toolbox Cognition Battery (NIHTB-CB), Delis-Kaplan Executive Function System (D-KEFS), and either the Wechsler Intelligence Scale for Children, 4^th^ Edition or the Wechsler Adult Intelligence Scale, 4^th^ Edition (WISC-IV and WAIS-IV, respectively), based on their age. Their parents also completed ratings using the Behavior Rating Inventory of Executive Function, 2^nd^ edition (BRIEF-2), To assess cognitive flexibility, the following tests were used: (1) NIHTB-CB Dimensional Change Card Sort Test (DCCS); (2) D-KEFS Verbal Fluency Test (VFT) Category Switching; (3) D-KEFS Trail Making Test (TMT) Number-Letter Switching; and (4) BRIEF-2 Shift. To assess inhibition, the following tests were used: (1) NIHTB-CB Flanker Attention & Inhibitory Control Test; (2) D-KEFS Color-Word Interference Test (CWIT) Inhibition; and (3) BRIEF-2 Inhibit. To assess working memory, the following tests were used: (1) NIHTB-CB List Sorting Working Memory; (2) WISC-IV or WAIS-IV Digit Span; (3) BRIEF-2 Working Memory.

### Statistical Analysis

The differences in flow metrices between CHD and healthy control groups were determined using Student’s T-Test. This was followed by quantifying differences in these CSF flow metrics between healthy controls and CHD subgrouping based on the type of heart defect at birth –Hypoplastic Left Heart Syndrome(HLHS), cyanotic, or left ventricular outflow tract obstruction (LVOTO) – using analysis of variance (ANOVA).

CSF flow metrics were then compared to CSF volumes with age at MRI and CHD status as covariates using multivariable regression. Lastly, a multivariable regression was performed comparing executive function outcomes with CSF flow metrics as primary exposure – and childhood opportunity index (COI) as a proxy for socio-demographic influence, and CHD status as covariates.

## Results

### Controls and CHD Group Description

There were no differences between the control and CHD groups based on age at MRI scan, age at neuropsychological testing, and gender (Table 1). There was also no difference in social-demographics factors including maternal education and COI between the two groups (Table 1). There was no difference in physical characteristics between groups (height and weight at time of MRI). Of the CHD patients, 10 were single ventricle physiology and 12 were bi-ventricular physiology (see Table 1 for a list of heart lesion sub-types).

**Table 1.**
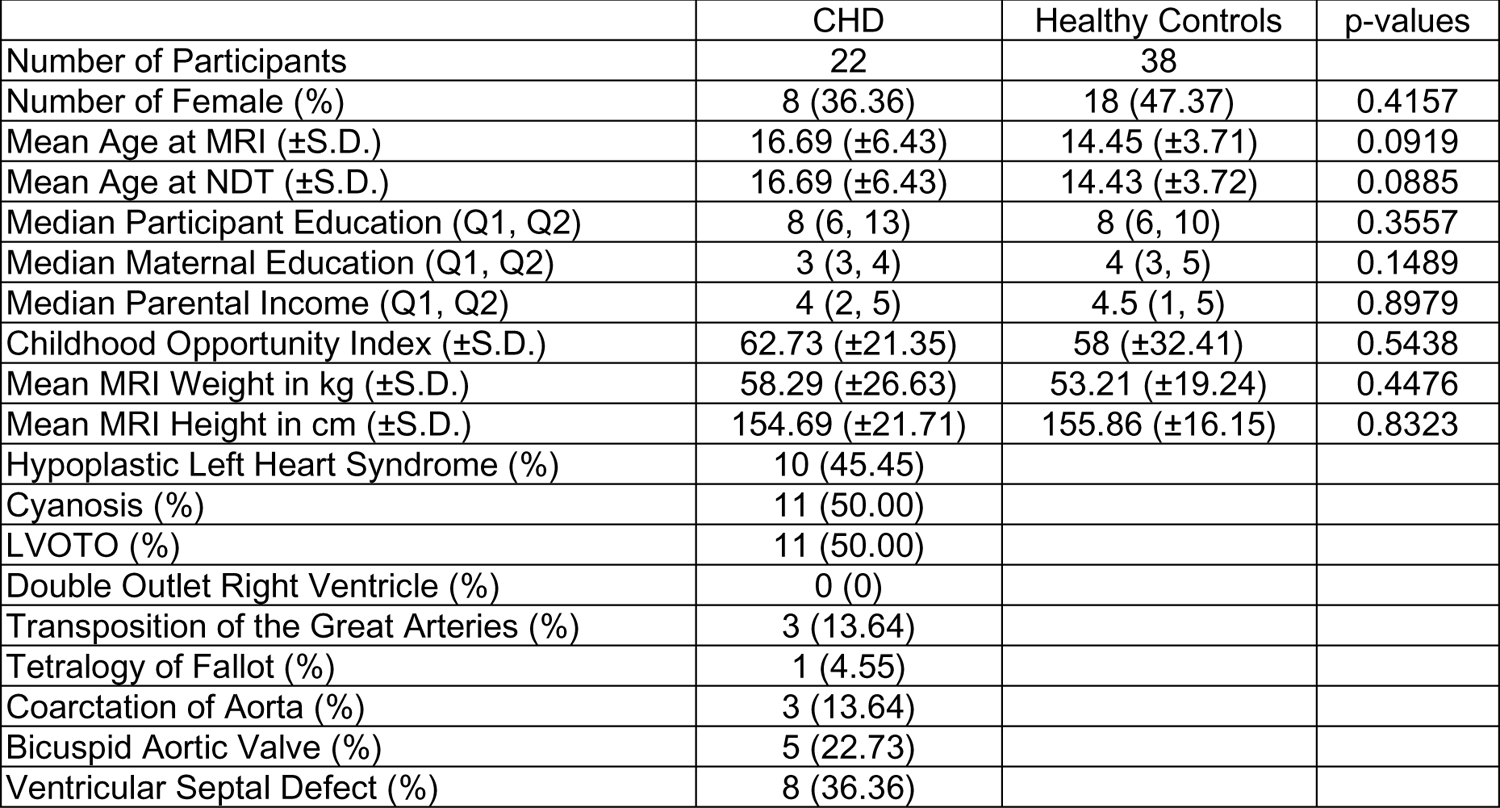
Demographic of the Groups and Cardiac Lesion Incidence.

|  | CHD | Healthy Controls | p-values |
| --- | --- | --- | --- |
| Number of Participants | 22 | 38 |  |
| Number of Female (%) | 8 (36.36) | 18 (47.37) | 0.4157 |
| Mean Age at MRI (±S.D.) | 16.69 (±6.43) | 14.45 (±3.71) | 0.0919 |
| Mean Age at NDT (±S.D.) | 16.69 (±6.43) | 14.43 (±3.72) | 0.0885 |
| Median Participant Education (Q1, Q2) | 8 (6, 13) | 8 (6, 10) | 0.3557 |
| Median Maternal Education (Q1, Q2) | 3 (3, 4) | 4 (3, 5) | 0.1489 |
| Median Parental Income (Q1, Q2) | 4 (2, 5) | 4.5 (1, 5) | 0.8979 |
| Childhood Opportunity Index (±S.D.) | 62.73 (±21.35) | 58 (±32.41) | 0.5438 |
| Mean MRI Weight in kg (±S.D.) | 58.29 (±26.63) | 53.21 (±19.24) | 0.4476 |
| Mean MRI Height in cm (±S.D.) | 154.69 (±21.71) | 155.86 (±16.15) | 0.8323 |
| Hypoplastic Left Heart Syndrome (%) | 10 (45.45) |  |  |
| Cyanosis (%) | 11 (50.00) |  |  |
| LVOTO (%) | 11 (50.00) |  |  |
| Double Outlet Right Ventricle (%) | 0 (0) |  |  |
| Transposition of the Great Arteries (%) | 3 (13.64) |  |  |
| Tetralogy of Fallot (%) | 1 (4.55) |  |  |
| Coarctation of Aorta (%) | 3 (13.64) |  |  |
| Bicuspid Aortic Valve (%) | 5 (22.73) |  |  |
| Ventricular Septal Defect (%) | 8 (36.36) |  |  |

### CSF Flow Metric Differences between Control and CHD groups

There were no differences in the following CSF flow metrics – APVel, RPVel, MVel, and VVel – between the control and CHD groups (Table 2) adjusted for age. However, the CHD group demonstrated higher overall Flow RMSD compared to controls (p=0.0353) (Table 2).

**Table 2:**
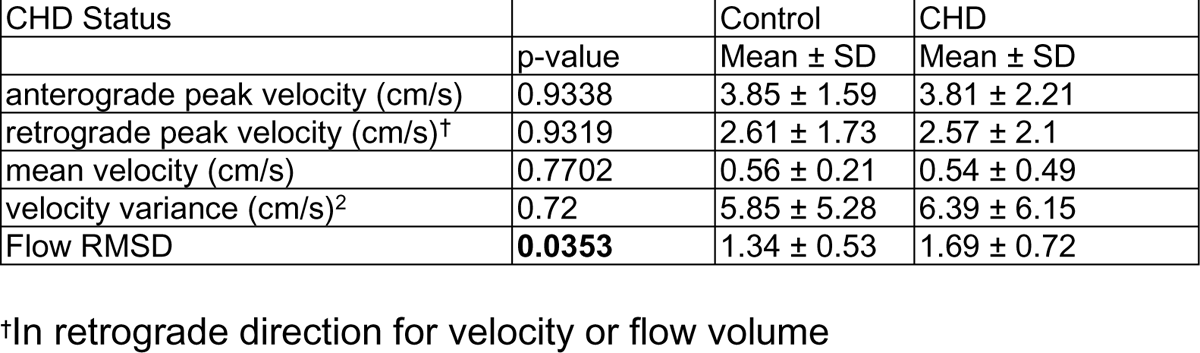
Differences in Flow Metrics Between CHD Participants and Healthy Controls.

We secondarily explored the univariate relationship between patient characteristics and CSF flow metrics (Supplementary Table 1). There were no differences in flow metrics between male and female participants in all groups combined. We did find that higher participant age was associated with lower CSF flow MVel (p=0.0224). Participant height and weight at MRI had no significant correlation with CSF flow metrics (Supplement Table 2a-b). We also explored the possibility of CSF flow metric differences between controls and different subtype of CHD (ANOVA) and did not see any significant differences (single ventricle physiology, cyanotic physiology, LVOTO anatomy) (Supplemental Table 3).

### Relationship of CSF Flow Metrics and CSF Volumes

In the multivariable analysis correlating CSF flow metrics to CSF volumes (Table 3), high APVel (p=0.0079) and high VVel (p=0.0109) were associated with greater third ventricle volume. In these same models, CHD group demonstrated greater third ventricle volume than controls adjusted for age.

**Table 3.** Association between CSF Flow and CSF Volume with Age at MRI and CHD status as covariates.

| CSF Volumes | CSF Flow Metric |  | CSF Flow |  | Age at MRI |  | CHD Status |  |
| --- | --- | --- | --- | --- | --- | --- | --- | --- |
|  |  | R-sq | p-value | direction | p-value | direction | p-value | direction |
| Left Lateral Ventricle | anterograde peak velocity | 0.0541 | 0.3701 | n.s. | 0.6343 | n.s. | 0.2069 | n.s. |
|  | retrograde peak velocity† | 0.0513 | 0.4221 | n.s. | 0.602 | n.s. | 0.2347 | n.s. |
|  | mean velocity | 0.0405 | 0.8399 | n.s. | 0.6812 | n.s. | 0.2233 | n.s. |
|  | velocity variance | 0.0399 | 0.9296 | n.s. | 0.5875 | n.s. | 0.2284 | n.s. |
|  | RMSD | 0.0444 | 0.6101 | n.s. | 0.5844 | n.s. | 0.1951 | n.s. |
| Left Inferior Lateral Ventricle | anterograde peak velocity | 0.1936 | 0.1797 | n.s. | 0.0402 | - | 0.0027 | CHD > Control |
|  | retrograde peak velocity† | 0.2237 | 0.0501 | n.s. | 0.0536 | n.s. | 0.0024 | CHD > Control |
|  | mean velocity | 0.1906 | 0.2055 | n.s. | 0.2065 | n.s. | 0.0086 | CHD > Control |
|  | velocity variance | 0.2 | 0.1357 | n.s. | 0.0534 | n.s. | 0.0037 | CHD > Control |
|  | RMSD | 0.1686 | 0.6839 | n.s. | 0.0538 | n.s. | 0.007 | CHD > Control |
| Right Lateral Ventricle | anterograde peak velocity | 0.0944 | 0.3433 | n.s. | 0.8682 | n.s. | 0.0336 | CHD > Control |
|  | retrograde peak velocity† | 0.0818 | 0.6933 | n.s. | 0.9182 | n.s. | 0.0404 | CHD > Control |
|  | mean velocity | 0.0797 | 0.8481 | n.s. | 0.8716 | n.s. | 0.0402 | CHD > Control |
|  | velocity variance | 0.0802 | 0.7989 | n.s. | 0.9328 | n.s. | 0.0403 | CHD > Control |
|  | RMSD | 0.0792 | 0.9313 | n.s. | 0.9286 | n.s. | 0.0448 | CHD > Control |
| Right Inferior Lateral Ventricle | anterograde peak velocity | 0.0719 | 0.4788 | n.s. | 0.121 | n.s. | 0.1292 | n.s. |
|  | retrograde peak velocity† | 0.0818 | 0.3001 | n.s. | 0.1401 | n.s. | 0.1305 | n.s. |
|  | mean velocity | 0.0643 | 0.806 | n.s. | 0.2076 | n.s. | 0.1649 | n.s. |
|  | velocity variance | 0.0801 | 0.3239 | n.s. | 0.1373 | n.s. | 0.1458 | n.s. |
|  | RMSD | 0.072 | 0.4767 | n.s. | 0.133 | n.s. | 0.2155 | n.s. |
| Third Ventricle | anterograde peak velocity | 0.2435 | **0.0079** | + | 0.2409 | n.s. | 0.0018 | CHD > Control |
|  | retrograde peak velocity† | 0.1921 | 0.0599 | n.s. | 0.3851 | n.s. | 0.0034 | CHD > Control |
|  | mean velocity | 0.1392 | 0.7022 | n.s. | 0.5089 | n.s. | 0.0076 | CHD > Control |
|  | velocity variance | 0.2353 | **0.0109** | + | 0.3689 | n.s. | 0.0039 | CHD > Control |
|  | RMSD | 0.1917 | 0.0608 | n.s. | 0.3504 | n.s. | 0.0203 | CHD > Control |
| Fourth Ventricle | anterograde peak velocity | 0.124 | 0.0582 | n.s. | 0.8745 | n.s. | 0.0592 | n.s. |
|  | retrograde peak velocity† | 0.0646 | 0.7819 | n.s. | 0.7561 | n.s. | 0.0837 | n.s. |
|  | mean velocity | 0.0669 | 0.6484 | n.s. | 0.9331 | n.s. | 0.0751 | n.s. |
|  | velocity variance | 0.0782 | 0.353 | n.s. | 0.7456 | n.s. | 0.0878 | n.s. |
|  | RMSD | 0.0684 | 0.5848 | n.s. | 0.7632 | n.s. | 0.1277 | n.s. |
| Extra-Axial CSF | anterograde peak velocity | 0.0508 | 0.2894 | n.s. | 0.5162 | n.s. | 0.2803 | n.s. |
|  | retrograde peak velocity† | 0.0303 | 0.8775 | n.s. | 0.5072 | n.s. | 0.3202 | n.s. |
|  | mean velocity | 0.0298 | 0.9783 | n.s. | 0.5346 | n.s. | 0.3375 | n.s. |
|  | velocity variance | 0.035 | 0.5999 | n.s. | 0.4786 | n.s. | 0.3236 | n.s. |
|  | RMSD | 0.0338 | 0.646 | n.s. | 0.4838 | n.s. | 0.3992 | n.s. |
| Whole Brain CSF | anterograde peak velocity | 0.0728 | 0.2184 | n.s. | 0.4446 | n.s. | 0.173 | n.s. |
|  | retrograde peak velocity† | 0.0454 | 0.9005 | n.s. | 0.4395 | n.s. | 0.2091 | n.s. |
|  | mean velocity | 0.0451 | 0.9937 | n.s. | 0.4748 | n.s. | 0.2209 | n.s. |
|  | velocity variance | 0.0518 | 0.5476 | n.s. | 0.4065 | n.s. | 0.2107 | n.s. |
|  | RMSD | 0.0493 | 0.6356 | n.s. | 0.4156 | n.s. | 0.2731 | n.s. |

†In the retrograde direction

### Relationship of CSF Flow Metrics and Neurocognitive Outcomes

We did not find any correlation between CSF metrics and either cognitive flexibility or working memory (Table 4). However, multiple correlations were found between particular CSF Flow metrics and inhibitory control. For example, Low APVel predicted poor performance on the NIHTB Flanker Attention & Inhibitory Control Test (p=0.0323), with no difference between control and CHD groups. Lower RMSD predicted worst performance on D-KEFS CWIT Inhibition (p=0.0181) participants with CHD demonstrated worse outcomes (p=0.0141) compared to controls (Table 4).

**Table 4.** CSF Flow Metric Predicting Outcomes with COI and CHD status as covariates.

|  |  |  | Independent |  | COI |  | CHD status |  |
| --- | --- | --- | --- | --- | --- | --- | --- | --- |
| Dependent | Independent | R<sup>2</sup> | p-value | direction | p-value | direction | p-value | direction |
| Working Memory |  |  |  |  |  |  |  |  |
| NIHTB-CB: List Sorting Working Memory | No Significant findings between Flow Metrics and Outcome |  |  |  |  |  |  |  |
| WISC-IV*: Digit Span | No Significant findings between Flow Metrics and Outcome |  |  |  |  |  |  |  |
| BRIEF-2 Parent Report: Working Memory | No Significant findings between Flow Metrics and Outcome |  |  |  |  |  |  |  |
| Cognitive Flexibility |  |  |  |  |  |  |  |  |
| NIHTB-CB: Dimensional Card Change Sort | No Significant findings between Flow Metrics and Outcome |  |  |  |  |  |  |  |
| D-KEFS TMT: Condition 4 – Number-Letter Switching | No Significant findings between Flow Metrics and Outcome |  |  |  |  |  |  |  |
| D-KEFS VFT: Condition 3 – Category Switching | No Significant findings between Flow Metrics and Outcome |  |  |  |  |  |  |  |
| D-KEFS VFT: Condition 3 – Correct Responses | No Significant findings between Flow Metrics and Outcome |  |  |  |  |  |  |  |
| BRIEF-2 Parent Report: Shift | No Significant findings between Flow Metrics and Outcome |  |  |  |  |  |  |  |
| Inhibition |  |  |  |  |  |  |  |  |
| NIHTB-CB: Flanker Attention & Inhibitory Control test | anterograde peak velocity (cm/s) | 0.0928 | **0.0323** | - | 0.9712 | n.s. | 0.2757 | n.s. |
| D-KEFS CWIT Inhibition | RMSD | 0.3161 | **0.0181** | + | **0.0348** | + | **0.0141** | CHD > Control |
| BRIEF-2 Parent Report: Inhibit | No Significant findings between Flow Metrics and Outcome |  |  |  |  |  |  |  |

## Discussion

Despite evidence associating structural CSF abnormalities with neurodevelopmental deficits in CHD[1,2], the relationship between abnormal CSF characteristics and CHD is not well understood and the potential impact of CSF flow properties on the CHD brain has not been explored previously. Thus, this is the first study to examine CSF flow properties in adolescents with CHD. In this study, we found that there were no differences in antegrade, retrograde or mean velocity between the two groups. However, while we did not see difference in velocity variances, we did detect a difference in RMSD, between the two groups. Importantly, RMSD models the CSF flow as a pulsatile phenomenon over the entire cardiac cycle, and we see that patients with CHD had higher variability in dynamic CSF flow, as measured by RMSD values, compared to controls. Taken together, while the whole CHD group exhibited higher dynamic CSF flow pulsatility compared to controls, the subset of CHD subjects with relatively reduced static and dynamic CSF flow pulsatility had the worst executive functioning, specifically the inhibition domain. These findings suggest that altered CSF flow pulsatility may be central to not only brain compensatory mechanisms but can also drive cognitive impairment in CHD. Further studies are needed to investigate possible mechanistic etiologies of aberrant CSF pulsatility (i.e. primary cardiac hemodynamic disturbances, intrinsic brain vascular stiffness, altered visco-elastic properties of tissue, or glial-lymphatic disturbances), which might result in evolving acquired small vessel disease (including microbleeds and white matter hyperintensities).

This finding of high RMSD variability in CSF flow of participants with CHD is likely a manifestation of the relationship between cardiopulmonary circulation and CSF flow observed in human and pre-clinical studies alike[14,28,29]. Due to the nature of their heart defect and surgical intervention, people with CHD tend to have a higher incidence of altered cardiac pulsations and cerebrovascular blood flow especially in complex forms of CHD[30]. They also have reduced compensatory tools to regulate cerebrovascular blood flow compared to healthy controls[31]. Since cardiopulmonary circulation, especially cerebral arterial and arteriole pulsations, contribute to CSF flow, alterations in this cerebrovascular blood flow commonly found in CHD might possibly explain some of this CSF flow variance as a direct effect of altered cardiopulmonary function. This could also explain the temporal variability in occurrence of peaks in the CSF flow cycle observed in the study cohort.

Another, related factor that could possibly contribute to this greater CSF flow variability over the cardiac cycle in CHD might be vessel compliance. The CHD population has a higher incidence as well as an earlier onset of arterial stiffness[32]. The stiffness of vessels could affect and impede the pulsatile wave forms from cardiopulmonary cycles transferring into the CSF circulation. Furthermore, fluid entering the CSF compartment via the choroid plexus relies on cerebral arterial pulsatility, while CSF follows a paravascular route as it traverses the brain parenchyma. This implies that both processes could be influenced by the compliance of these vessels, particularly the pial and penetrating arteries of the brain. Additionally, the CSF route through the brain parenchyma, and thus CSF flow variability, could be affected by brain tissue compliance. Brain tissue compliance is partly determined by white matter density and myelination which are sometimes reduced in CHD. Lastly, glymphatic clearance mechanism, which are still elusive, could exert indirect effect on the CSF flow variability through alterations to hydrostatic pressure downstream of CSF flow[33].

Another direct effect on CSF flow variability over the cardiac cycle in CHD might be CSF compartment size. While we did not see a significant association between RMSD values and Ventricular size, our results showed that both anterograde peak velocity and velocity variance were greater in participants with larger third ventricle volume. CSF compartment size could alter the CSF flow rate, as demonstrated by Longatti and colleagues in the mechanical model of third and fourth ventricle sizes and the relationship to the flow rate across the Aqueduct of Sylvius[34]. This then begs the question of what factors might be affecting third ventricular volume which points to possible hypothalamic and other subcortical involvement. These regional structures could be reduced in CHD. Choroid plexus function could also impact CSF flow rate, and our prior study in neonates indicate a higher incidence of abnormal choroid plexus structures in CHD. While choroid plexus function was beyond the scope of this study, choroid plexus function assessment should be explored in conjunction with CSF features in further studies.

In terms of outcomes, we observed significant association between CSF flow characteristics and inhibitory control. Decreased mean velocity predicted poor inhibition on testing from the NIHTB, while increased variability in pulsatile CSF flow, as measured by RMSD, predicted better performance on testing from the D-KEFS. Recall that mean velocity measure for each participant is calculated from the average of the twenty velocity time points over the entire flow cycle. Lower mean velocity is an interplay of balance between flow velocities in both anterograde and retrograde velocities. This result with testing from the NIHTB seems to indicate that participants with anterograde velocities comparable in magnitude to the retrograde velocities – in other words participants with averaging of flow velocities closer to zero – tend to have worse inhibitory performance. In the model between RMSD and D-KEFS CWIT Inhibition, the higher RMSD had better outcome. This implies that participants with higher variability in pulsatile flow over the cardiac cycle performed better on this test. This suggests the greater pulsatile variability might be associated with some type of compensatory mechanism.

These findings of selective vulnerability of inhibitory control compared to other executive functioning domains raises intriguing questions regarding CSF flow and inhibition issues in non-CHD contexts. Exploring the neural mechanisms underlying this phenomenon could offer insights into potential avenues for risk stratification among CHD patients. Identifying individuals who may benefit from interventions targeting inhibition, through CSF flow features, could be a crucial step in improving their outcomes. By delving deeper into the interplay between CSF dynamics, neural mechanisms, and executive function, we may uncover novel strategies to optimize interventions and improve outcomes for CHD patients.

Due to it being the first study of its kind in CHD population, there were unanticipated challenges and limitations that arose, which could serve as lessons to inform and improve further studies. While the MRI sequence used to CSF flow measurement was cardiac gated, the limits of technology and data collection prevented acquiring EKG signal which could have potentially been used to assess the correlation between cardiac rhythm and the overall pulsatile flow. In future studies, the acquisition of the cardiac signal and comparison to the overall CSF flow is necessary. We also observed wide variation in the occurrence of anterograde and retrograde peak velocities between participants in both controls and CHD groups. This suggests there may be a phase differential component that might be interrelated to cardiac MR dynamic metrics and the triggering of the MRI sequence. However, without EKG or specific cardiac MRI hemodynamic parameters, it was not possible to further examine this possible phase difference phenomenon. To mitigate this possible confound, for our analysis, we aligned the CSF flow by the anterograde peak velocity in the RMSD calculations.

This project is also the first to study CSF flow at not only at the individual level with isolated measurements but also by comprehensively analyzing it as a pulsatile phenomenon across the cardiac cycle at the group level. Furthermore, as part of our study, we developed an adaptable and comprehensive PCMRI processing pipeline in MATLAB – that allows repeatability through use of savable and transferable masks – that can capture additional features that might be important to define CSF flow abnormalities.

We fashioned our initial analysis to be similar to the validated measurements used in prior CSF flow studies. Namely, we examined the standard features and measurements of CSF flow typically available on commercial software such as average or peak velocities. However, these static features of average and peak velocities showed no statistical difference between the CHD group and the control group. This lack of difference between the CHD and control groups is partly explained by the nature of these standard measurements that collapse high-dimensionality signal data into single values, which are incapable of distinguishing the groups due to the highly variable nature of CSF flow in the CHD group.

## CONCLUSION

Taken together, while the whole CHD group exhibited higher dynamic CSF flow pulsatility compared to controls, the subset of CHD subjects with relatively reduced static and dynamic CSF flow pulsatility had the worst executive functioning, specifically the inhibition domain. These findings suggest that altered CSF flow pulsatility may be central to not only brain compensatory mechanisms but can also drive cognitive impairment in CHD. Further studies are needed to investigate possible mechanistic etiologies of aberrant CSF pulsatility (i.e. primary cardiac hemodynamic disturbances, intrinsic brain vascular stiffness, altered visco-elastic properties of tissue, or glial-lymphatic disturbances). Future work is also needed to examine the relationship between CSF pulsatility characteristics and evolving acquired small vessel brain injury disease (including microbleeds and white matter hyperintensities) that are known to be present in CHD. The third ventricular volume association with certain CSF flow features also raises the possibility of mechanical effects of volume compartments and tissue compliance on flow and is worth further consideration in characterizing the clearance function of CSF and the relation to dysmaturation of subcortical structure, also known to be present in CHD. In-depth studies characterizing the factors that drive the pulsatility of CSF flow are essential for greater insights into the intricate mechanisms underlying CSF circulation and its link to CHD-related brain health outcomes.

## Data Availability

Individual participant data that underlie the results reported in this article, after deidentification (text, tables, figures, and appendices), Study Protocol, and Statistical Analysis Plan, are available upon formal request. Requests should be submitted to the corresponding author.

## References

1. Panigrahy, A.; Lee, V.; Ceschin, R.; Zuccoli, G.; Beluk, N.; Khalifa, O.; Votava-Smith, J.K.; DeBrunner, M.; Munoz, R.; Domnina, Y. Brain dysplasia associated with ciliary dysfunction in infants with congenital heart disease. The Journal of pediatrics 2016, 178, 141–148. e141.

2. Heye, K.N.; Knirsch, W.; Latal, B.; Scheer, I.; Wetterling, K.; Hahn, A.; Akintürk, H.; Schranz, D.; Beck, I.; Reich, B. Reduction of brain volumes after neonatal cardiopulmonary bypass surgery in single-ventricle congenital heart disease before Fontan completion. Pediatric research 2018, 83, 63–70.

3. Reich, B.; Heye, K.N.; Tuura, R.O.G.; Beck, I.; Wetterling, K.; Hahn, A.; Aktintürk, H.; Schranz, D.; Jux, C.; Kretschmar, O. Interrelationship between hemodynamics, brain volumes, and outcome in hypoplastic left heart syndrome. The Annals of thoracic surgery 2019, 107, 1838–1844.

4. Kelly, C.J.; Makropoulos, A.; Cordero-Grande, L.; Hutter, J.; Price, A.; Hughes, E.; Murgasova, M.; Teixeira, R.P.A.; Steinweg, J.K.; Kulkarni, S. Impaired development of the cerebral cortex in infants with congenital heart disease is correlated to reduced cerebral oxygen delivery. Scientific reports 2017, 7, 1–10.

5. Ng, I.H.; Bonthrone, A.F.; Kelly, C.J.; Cordero-Grande, L.; Hughes, E.J.; Price, A.N.; Hutter, J.; Victor, S.; Schuh, A.; Rueckert, D. Investigating altered brain development in infants with congenital heart disease using tensor-based morphometry. Scientific reports 2020, 10, 1–10.

6. Ceschin, R.; Wisnowski, J.L.; Paquette, L.B.; Nelson, M.D.; Blüml, S.; Panigrahy, A. Developmental synergy between thalamic structure and interhemispheric connectivity in the visual system of preterm infants. NeuroImage: Clinical 2015, 8, 462–472.

7. Glauser, T.A.; Rorke, L.B.; Weinberg, P.M.; Clancy, R.R. Congenital brain anomalies associated with the hypoplastic left heart syndrome. Pediatrics 1990, 85, 984–990.

8. Brossard-Racine, M.; Du Plessis, A.J.; Vezina, G.; Robertson, R.; Bulas, D.; Evangelou, I.E.; Donofrio, M.; Freeman, D.; Limperopoulos, C. Prevalence and spectrum of in utero structural brain abnormalities in fetuses with complex congenital heart disease. American Journal of Neuroradiology 2014, 35, 1593–1599.

9. Panigrahy, A.; Ceschin, R.; Lee, V.; Beluk, N.; Khalifa, O.; Zuccoli, G.; Munoz, R.; Domnina, Y.; Wearden, P.; Morell, V. Respiratory Ciliary Motion Defect Predict Regional Brain Abnormalities and Increased Extra Axial CSF Fluid in Neonates With Complex Congenital Heart Disease. Circulation 2014, 130, A16570–A16570.

10. Owen, M.; Shevell, M.; Donofrio, M.; Majnemer, A.; McCarter, R.; Vezina, G.; Bouyssi-Kobar, M.; Evangelou, I.; Freeman, D.; Weisenfeld, N. Brain volume and neurobehavior in newborns with complex congenital heart defects. The Journal of pediatrics 2014, 164, 1121–1127. e1121.

11. Panigrahy, A.; Votava-Smith, J.; Lee, V.; Gabriel, G.; Klena, N.; Gibbs, B.; Reynolds, W.T.; Zuccoli, G.; O’Neil, S.; Schmithorst, V.J. Abnormal Brain Connectivity and Poor Neurodevelopmental Outcome in Congenital Heart Disease Patients With Subtle Brain Dysplasia. Circulation 2015, 132, A16541–A16541.

12. Ueno, H. MOTOR PROTEINS OF CILIA. Integrated Nano-Biomechanics 2018, 155.

13. Louvi, A.; Grove, E.A. Cilia in the CNS: the quiet organelle claims center stage. Neuron 2011, 69, 1046–1060.

14. Iliff, J.J.; Wang, M.; Liao, Y.; Plogg, B.A.; Peng, W.; Gundersen, G.A.; Benveniste, H.; Vates, G.E.; Deane, R.; Goldman, S.A. A paravascular pathway facilitates CSF flow through the brain parenchyma and the clearance of interstitial solutes, including amyloid β. Science translational medicine 2012, 4, 147ra111–147ra111.

15. Battal, B.; Kocaoglu, M.; Bulakbasi, N.; Husmen, G.; Tuba Sanal, H.; Tayfun, C. Cerebrospinal fluid flow imaging by using phase-contrast MR technique. The British journal of radiology 2011, 84, 758–765.

16. Mestre, H.; Tithof, J.; Du, T.; Song, W.; Peng, W.; Sweeney, A.M.; Olveda, G.; Thomas, J.H.; Nedergaard, M.; Kelley, D.H. Flow of cerebrospinal fluid is driven by arterial pulsations and is reduced in hypertension. Nature communications 2018, 9, 1–9.

17. Shen, M.D. Cerebrospinal fluid and the early brain development of autism. Journal of neurodevelopmental disorders 2018, 10, 1–10.

18. Attier-Zmudka, J.; Sérot, J.-M.; Valluy, J.; Saffarini, M.; Macaret, A.-S.; Diouf, M.; Dao, S.; Douadi, Y.; Malinowski, K.P.; Balédent, O. Decreased cerebrospinal fluid flow is associated with cognitive deficit in elderly patients. Frontiers in aging neuroscience 2019, 11, 87.

19. Sahel, A.; Ceschin, R.; Badaly, D.; Lewis, M.; Lee, V.K.; Wallace, J.; Weinberg, J.; Schmithorst, V.; Lo, C.; Panigrahy, A. Increased Cerebello-Prefrontal Connectivity Predicts Poor Executive Function in Congenital Heart Disease. Journal of Clinical Medicine 2023, 12, 5264.

20. Wallace, J.; Ceschin, R.; Lee, V.K.; Beluk, N.H.; Burns, C.; Beers, S.; Lo, C.; Panigrahy, A.; Badaly, D. Psychometric Properties of the NIH Toolbox Cognition and Emotion Batteries Among Children and Adolescents with Congenital Heart Defects. *medRxiv* 2023.

21. Schmithorst, V.J.; Adams, P.S.; Badaly, D.; Lee, V.K.; Wallace, J.; Beluk, N.; Votava-Smith, J.K.; Weinberg, J.G.; Beers, S.R.; Detterich, J. Impaired Neurovascular Function Underlies Poor Neurocognitive Outcomes and Is Associated with Nitric Oxide Bioavailability in Congenital Heart Disease. Metabolites 2022, 12, 882.

22. Schmithorst, V.J.; Badaly, D.; Beers, S.R.; Lee, V.K.; Weinberg, J.; Lo, C.W.; Panigrahy, A. Relationships between regional cerebral blood flow and neurocognitive outcomes in children and adolescents with congenital heart disease. In Proceedings of the Seminars in Thoracic and Cardiovascular Surgery, 2022; pp. 1285–1295.

23. Badaly, D.; Beers, S.R.; Ceschin, R.; Lee, V.K.; Sulaiman, S.; Zahner, A.; Wallace, J.; Berdaa-Sahel, A.; Burns, C.; Lo, C.W. Cerebellar and prefrontal structures associated with executive functioning in pediatric patients with congenital heart defects. Frontiers in Neurology 2022, 13, 827780.

24. Desikan, R.S.; Ségonne, F.; Fischl, B.; Quinn, B.T.; Dickerson, B.C.; Blacker, D.; Buckner, R.L.; Dale, A.M.; Maguire, R.P.; Hyman, B.T. An automated labeling system for subdividing the human cerebral cortex on MRI scans into gyral based regions of interest. Neuroimage 2006, 31, 968–980.

25. Reuter, M.; Rosas, H.D.; Fischl, B. Highly accurate inverse consistent registration: a robust approach. Neuroimage 2010, 53, 1181–1196.

26. Zhang, Y.; Brady, M.; Smith, S. Segmentation of brain MR images through a hidden Markov random field model and the expectation-maximization algorithm. IEEE transactions on medical imaging 2001, 20, 45–57.

27. Diamond, A. Executive functions. Annual review of psychology 2013, 64, 135–168.

28. Bilston, L.E.; Stoodley, M.A.; Fletcher, D.F. The influence of the relative timing of arterial and subarachnoid space pulse waves on spinal perivascular cerebrospinal fluid flow as a possible factor in syrinx development. Journal of neurosurgery 2010, 112, 808–813.

29. Schroth, G.; Klose, U. Cerebrospinal fluid flow: I. Physiology of cardiac-related pulsation. Neuroradiology 1992, 35, 1–9.

30. De Silvestro, A.; Natalucci, G.; Feldmann, M.; Hagmann, C.; Nguyen, T.D.; Coraj, S.; Jakab, A.; Kottke, R.; Latal, B.; Knirsch, W. Effects of hemodynamic alterations and oxygen saturation on cerebral perfusion in congenital heart disease. Pediatric Research 2024, 1–9.

31. Donofrio, M.; Bremer, Y.; Schieken, R.; Gennings, C.; Morton, L.; Eidem, B.; Cetta, F.; Falkensammer, C.; Huhta, J.; Kleinman, C. Autoregulation of cerebral blood flow in fetuses with congenital heart disease: the brain sparing effect. Pediatric cardiology 2003, 24, 436–443.

32. Sandhu, K.; Pepe, S.; Smolich, J.J.; Cheung, M.M.; Mynard, J.P. Arterial stiffness in congenital heart disease. Heart, Lung and Circulation 2021, 30, 1602–1612.

33. Jessen, N.A.; Munk, A.S.F.; Lundgaard, I.; Nedergaard, M. The glymphatic system: a beginner’s guide. Neurochemical research 2015, 40, 2583–2599.

34. Longatti, P.; Fiorindi, A.; Peruzzo, P.; Basaldella, L.; Susin, F.M. Form follows function: estimation of CSF flow in the third ventricle–aqueduct–fourth ventricle complex modeled as a diffuser/nozzle pump. Journal of neurosurgery 2019, 133, 894–901.

